# Using Mendelian Randomization to model the causal effect of cancer on health economic outcomes and to simulate the cost-effectiveness of anti-cancer interventions

**DOI:** 10.1101/2023.02.06.23285521

**Authors:** Padraig Dixon, Richard M Martin, Sean Harrison

**Author notes:** Corresponding author. Padraig Dixon, Nuffield Department of Primary Care Health Sciences, University of Oxford, Radcliffe Primary Care Building, Radcliffe Observatory Quarter, Woodstock Rd, Oxford OX2 6GG. These authors contributed equally (Padraig Dixon, Sean Harrison).

## Abstract

**BACKGROUND:** Cancer is associated with significant economic impacts. Quantifying the scale of these impacts is challenged by confounding variables that jointly influence both cancer status and economic outcomes such as healthcare costs and quality of life. Moreover, the increasing costs attributed to cancer drug development complicate the cost-effective provision of cancer care.

**METHODS:** We address both challenges in this paper by using germline genetic variation in the risk of incident cancer as instrumental variables in Mendelian Randomization analyses of eight cancers. We developed causal estimates of the genetically predicted effect of bladder, breast, colorectal, lung, multiple myeloma, ovarian, prostate and thyroid cancers on healthcare costs and quality adjusted life years (QALYs) using outcome data drawn from the UK Biobank cohort. We then used Mendelian Randomization to model a hypothetical population-wide preventative intervention based on a repurposed class of anti-diabetic drugs known as sodium-glucose co-transporter-2 (SGLT2) inhibitors very recently shown to reduce the odds of incident prostate cancer.

**RESULTS:** Genetic liability to prostate cancer and to breast cancer had material causal impacts on healthcare costs and QALYs. Mendelian Randomization results for the less common cancers were associated with considerable uncertainty. SGLT2 inhibition was unlikely to be a cost-effective preventative intervention for prostate cancer, although this conclusion depended on the price at which these drugs would be offered for a novel anti-cancer indication.

**IMPLICATIONS:** Our new causal estimates of cancer exposures on health economic outcomes may be used as inputs into decision analytic models of cancer interventions such as screening programmes or simulations of longer-term outcomes associated with therapies investigated in RCTs with short follow-ups. Our new method allows us to rapidly and efficiently estimate the cost-effectiveness of a hypothetical population-scale anti-cancer intervention to inform and complement other means of assessing long-term intervention cost-effectiveness.

## 1 Introduction

Cancer, a major global cause of morbidity and death (1-4), is associated with important economic impacts on patients, carers and health systems (5-8). Increases in the average age of populations, and improvements in the detection and treatment of cancer, are creating growing cohorts of individuals who have received cancer treatment, who are receiving treatment, or who may receive some form of cancer intervention for the rest of their lives (6, 9, 10). In turn, extended treatment modalities, new adjuvant regimes and increases in the costs of therapies, challenge the cost-effective delivery of care for cancer patients (8, 11-16).

Knowledge of how cancer status affects health economic outcomes such as healthcare cost and patient quality of life is central to providing cost-effective care to patients. Conventional study designs that examine associations between cancer status and these outcomes can be affected by measurement error and by omitted variable bias. Any traits, behaviours, or disease processes (including prodromal processes) that influence or are influenced by cancer status are likely in most cases to independently influence healthcare costs, quality of life or both.

For example, risk factors such as cigarette smoking or adverse weight profiles may co-occur, are known to independently and jointly influence risk for many cancers (17-21), and have quantitatively important causal influences on health economic outcomes (22-25). Cohort studies (for example (26)) that attribute costs to cancer patients may account for direct costs following a diagnosis of cancer, but cannot reflect all causal impacts associated with liability to cancer or cancer diagnoses because in general it is not possible to identify all confounding variables, to assess their contribution without measurement error, to condition away their influence in quantitative analysis, or to include all downstream causal impacts.

A further challenge to providing affordable care relates to the high and increasing costs associated with anti-cancer pharmacotherapies (27). The inflation-adjusted launch price of newly approved cancer drugs in the United States increased at 10% every year between 1995 and 2013 (28), implying a doubling of prices roughly every 7 years, and amounting to average annual increases of $8,000. Annual average costs for novel anti-cancer drugs in the US are now over $100,000 and considerably more where such drugs are taken in combination (13, 29, 30). This is despite weak or no evidence of a connection between high prices, development costs and therapeutic efficacy for drugs as a whole (31-34). These challenges are not confined to any one country (8, 14, 15, 35-38) and serve to emphasize the formidable costs associated with developing and remunerating anti-cancer pharmacotherapies.

Identifying cost-effective cancer therapies, and improving the efficiency of drug development, therefore offers considerable promise for improving the value of cancer care. This paper addresses both challenges. We report the first use of Mendelian Randomization (39, 40) to estimate the causal effect of genetic liability to different site-specific cancers on, respectively, healthcare cost and quality of life.

Mendelian Randomization (39-43) is a type of instrumental variable analysis that relies on the natural experiment of quasi-random allocation of genetic variation from parents to children at conception. Some of this genetic variation is known to influence susceptibility to cancer and is – in principle, and with assumptions that we describe below – independent of post-conception influences that would otherwise confound the relationship between cancer status and health economic outcomes in conventional analytic modelling. We then use these Mendelian Randomization estimates to efficiently assess the cost-effectiveness of a novel and potentially affordable population-wide anti-cancer prophylactic drug therapy repurposed from the treatment of diabetes.

Together, these two work strands demonstrate the feasibility of using Mendelian Randomization to study the impact of cancer exposures on health economic outcomes, as well as the practicality of these methods to rapidly and efficiently estimate the cost-effectiveness of anti-cancer interventions to complement, prioritise or inform the design of randomized controlled trials or other evaluations of cancer therapies.

## 2 Methods

### 2.1 Introduction

We used Mendelian Randomization to estimate the causal effect of cancer on healthcare costs and on quality of life, and in turn, quality adjusted life years. Many introductions to Mendelian Randomization are available (39, 41-43). Briefly, Mendelian Randomization relies on Mendel’s first (random segregation of alleles at conception) and second (independent assortment of alleles) laws of inheritance, which describe how genetic variants are acquired by children from their parents. An allele refers to the specific form of genetic variation found a particular location in the genome.

Some of these genetic variants transmitted in this conditionally random manner are known to influence the risk of expressing specific phenotypes, including disease phenotypes such as cancer. At a population level, the association of genetic variants that influence the risk of, for example, a site-specific cancer, permits the statistical identification of groups that differ in the levels of their exposure to the risk for that cancer.

Genetic variants inherited according to Mendelian principles are therefore strong candidates as instrumental variables since their association with an exposure of interest can – in principle – be demonstrated (ideally from genome wide association studies), thus satisfying the first assumption (“relevance”) of valid instrumental variable analysis.

The second assumption necessary for valid instrumental variable analysis is that the instrument be independent of all confounding variables; put differently, that the instrument should be as “good as randomly assigned” across different groups defined by confounding variables. This assumption cannot be tested, as this requirement encompasses all manner of confounding variables, including those are unmeasured, unobserved, and unknown. However, the assignment of genetic variants at conception means that postnatal influences are unlikely to confound associations of interest, meaning that differences in outcomes between groups defined by genetic liability to cancer (in this case) may be more causally attributed to the cancer exposure itself.

The third assumption for valid instrumental variable analysis, the exclusion restriction, requires that the instrument influence the outcome only via the exposure of interest. This is generally untestable since it implies independence of the instrument from all outcomes (including potential outcomes) conditional on the exposure. Moreover, genetic instrument variables of the type used in Mendelian Randomization are very likely to violate this assumption in two ways.

The first potential violation is linkage disequilibrium, which refers to correlation between genetic variants which may arise when they are located near to each other on the genome. Linkage disequilibrium will violate the exclusion restriction if correlated variants affect the outcome other than via the exposure of interest. The second potential violation of the exclusion restriction may arise due to pleiotropy, which refers to the effect of a variant on more than one phenotype. Pleiotropy will violate the exclusion restriction if these other phenotypes also affect the outcome through channels that operate independently of the exposure.

Below, we describe how we identified genetic variants to serve as instrumental variables for as wide a variety of site-specific cancers as possible in our Mendelian Randomization analysis. We also describe the steps taken to ensure our results were robust to potential violations of the instrumental variable assumptions.

### 2.2 Polygenic risk score instrumental variable models

We developed polygenic risk scores (PRSs) for each cancer, the specific steps in the construction of which are described in detail below. The PRSs, which are functions of the weighted log-odds of developing each specific cancer, were used as instruments in “just identified” two-stage-least square (2SLS) instrumental variable models. “Just identified” models have as many instruments as there are exposures whereas “over identified” models have more instruments than exposures. We used the *ivreg2* package in Stata (version 17.0) with robust standard errors to estimate these models.

In the first stage of the 2SLS model for each cancer, a cancer exposure variable was regressed on the PRSs with age at baseline assessment, sex, 40 genetic principal components (to control for the effects of ancestry-related population structure), and UK Biobank recruitment centre as covariates. The predicted values of the coefficient on the instrument were then used in the second stage regressions with either costs or quality of life as the outcomes. F-statistics from the first stage regressions were inspected to assess instrument strength to assess whether the PRS for each cancer was sufficiently associated with the observed cancers in the population (23).

Our Mendelian randomization analysis estimates the mean difference in the outcomes using an additive structural mean model (24–26), interpreted as the average change in each outcome caused by the hypothetical average effect of having the cancer versus not across the population (44). This assumes that a constant effect of genetic liability to each cancer on the cost and quality of life outcomes. An alternative assumption is to interpret the results as a local average treatment effect, which assumes a monotonic effect of genetic variants on each cancer exposure (45).

We compared the Mendelian randomization estimates with estimates from conventional separate multivariable linear regressions for QALYs and healthcare costs, with age, sex, 40 genetic principal components, and recruitment centre as covariates. We performed Hausman endogeneity tests (27), in which a small p-value indicates there was evidence the Mendelian Randomization and multivariable effect estimates were different, although the power of these tests is relatively low (46).

### 2.3 Sensitivity analyses

We conducted a variety of sensitivity analyses. We performed a multivariable adjusted analysis with additional variables (Townsend Deprivation Index, household income, highest educational qualification, ever smoked, and body mass index), to reduce the risk of confounding in the multivariable adjusted analyses.

We re-ran the main Mendelian Randomization analysis stratified by sex and age group (less than 50 years, 50 to 59 years, and 60+ years), both separately and together. We also stratified by when the participant was diagnosed with cancer, either before or after recruitment into UK Biobank, as participants who received their cancer diagnosis before recruitment were more likely to have smaller healthcare costs associated with diagnosis, and possibly curative treatment, but more likely to have greater healthcare costs associated with managing progressing cancers, and vice versa for participants who received their cancer diagnosis after recruitment.

We also examined potential violations of the exclusion restriction due to horizontal pleiotropy. This occurs when a variant influences the outcome through a channel other than the cancer of interest. This may be via another disease, trait, behaviour, or health condition other than the cancer specifically under investigation.

To test for the presence of pleiotropy, we estimated over-identified Mendelian Randomization models comprising the individual SNPs used in constructing cancer-specific PRSs. Heterogeneity in the effect of these SNPs on the cost and quality of life outcomes could indicate a violation of the exclusion restriction.

Heterogeneity in effect estimates across SNPs can be assessed by comparing Cochran’s Q statistic 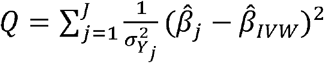 to the critical values of a chi-squared distribution. In this formula, there are *J* total SNPs, 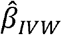 is the inverse variance weighted (IVW) effect calculated for all *J* SNPs, 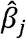 is the effect estimate for a specific SNP *j*, and 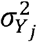 the variance of the SNP-outcome association. The intuition for this formula is that any 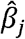 with a large impact on the outcome that different from the overall IVW effect of all SNPs on the outcome may suggest multiple pleiotropic channels of influence of that SNP on the outcome not mediated by the specific cancer exposure investigated.

We used consensus methods and a modelling method (47) for our sensitivity analyses, each of which embodied different types of assumption about whether and in which respects the exclusion restriction may be violated. We first estimated the IVW estimates needed to calculate Cochran’s Q statistic, which involved calculating instrumental variable estimates for each SNP separately, and then combining them in random-effects meta-analysis with weights determined by the precision of the association between the SNP and the outcomes (healthcare costs and quality of life respectively). This assumes no violations of the exclusion restriction or no net effect of any such violations on the point estimates.

We then compared this restrictive baseline model against more models that are robust to different types of exclusion restriction violation. We implemented the Mendelian Randomization Egger estimator, which is consistent even if all SNPs are pleiotropic provided additional assumptions (relating to the association between instrument strength and the direct pleiotropic effect of SNPs) are met – see (48, 49). We also implemented a penalized weighted median estimator, which is consistent if at least 50% if the SNPs are valid instrumental variables (50). We also implemented a weighted mode estimator, which is consistent if the largest homogenous cluster of SNPs are valid even if more than 50% of SNPs are invalid as instrumental variables (51).

Note that results from these estimators are on a different scale to the instrumental variable estimates obtained from the polygenic risk score models. The over-identified sensitivity analyses reflect the measurement scales used for the outcomes in the respective source genome-wide association studies (GWASs), which are based on logistic regression. The scale of these outcomes therefore reflects a change in costs or quality of life per unit change in the log-odds of cancer status. These results are on a relative rather than absolute scale, reflecting the relative increase in genetic liability to cancer from increasing the number of risk-increasing alleles. By contrast, the effect estimates obtained from the 2SLS just-identified models are estimates of genetically influenced changes in disease status in the analysis population.

### 2.4 Modelling the cost-effectiveness of a novel anti-cancer intervention

We used our new estimates of the causal impact of cancer on healthcare cost and QALYs to simulate the impact of a new population-wide anti-cancer intervention. Zheng et al (52) used a Mendelian Randomization study design to assess the impact of sodium-glucose cotransporter 2 (SGLT2) inhibitors (a type of oral glucose lowering drug) on prostate cancer risk. Their analysis found that SGLT2 inhibition, equivalent to a 1.09% reduction in HbA1c levels, lowered the odds of prostate cancer by 71% (OR=0.29, 95% CI: 0.13 to 0.65) in their primary analysis and by 49% in replication analysis (OR=0.51, 95% CI: 0.33 to 0.79).

In the absence of data on how such the hypothetical SGLT2 intervention would be dosed and costed for this new indication, we assumed that it would be offered in the form of 300mg of canagliflozin, a particular type of SGLT2 inhibitor. As a monotherapy, a dose of 300mg of canagliflozin had a broadly similar (53, 54) impact on HbA1c (approximately a 1% reduction) as did the MR estimates in the Zheng et al (52) paper. These comparisons are necessarily rather coarse since the Zheng et al (52) estimates relate to a largely unselected population, while the effectiveness estimates in the trials relate to effectiveness amongst diabetic patients, are dependent (to some degree) on baseline HbA1c levels, and we assume in our analysis no side-effects from prophylactic treatment with these inhibitors.

We further note that the Zheng et al (52) estimates pertain to lifelong SGLT2 inhibition, and initiation of daily canagliflozin by, for example, the middle to early old age men represented in our UK Biobank cohort will likely understate the cost-effectiveness of this intervention. These are restrictive assumptions that may not apply to the use of these drugs in a “real-world” deployment, but at least serve to indicate how these methodologies can rapidly provide an initial estimate of cost-effectiveness for a novel population-wide anti-cancer intervention.

Our hypothetical intervention therefore assumed that every man in the analysis sample was offered and took daily SGLT2 inhibitors (and specifically 300mg of canagliflozin) from their recruitment into the UK Biobank cohort. To estimate the causal impact of prostate cancer on healthcare cost and QALYs, we needed to estimate 3 quantities: 1) the proportion of men who developed prostate cancer, 2) the proportion of men who wouldn’t have developed prostate cancer had they received the intervention, and 3) the total healthcare and QALY cost saved by preventing those men from developing prostate cancer. Each of these three quantities had associated statistical imprecision.

For the first quantity, we identified the proportion of all men in UK Biobank who were diagnosed with prostate cancer, as well as within different age bands (less than 50 years, 50 to 59 years, and 60+ years), estimating the relevant standard errors (SE) of each proportion:

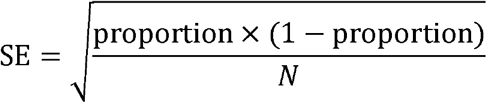

where N is the total number of men (overall, or within each age band).

For the second quantity, we converted the odds ratios from Zheng et al (52) and the proportions estimated in quantity 1 into risk differences:

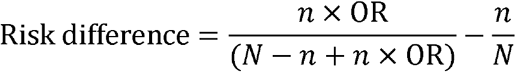

where *n* is the number of men with prostate cancer, *N* is the total number of men, and OR is the odds ratio. For example, 6,155 of 144,032 men (4.3%) in UK Biobank were diagnosed with prostate cancer up to 31 March 2017, so an OR of 0.29 from Zheng et al (52) gives a risk difference of -3.0%.

For the third quantity, we used the relevant multivariable adjusted or Mendelian randomization results estimated in section 2.2, both for total annual healthcare costs and QALYs.

We multiplied the risk difference (quantity 2) by the relevant multivariable adjusted or Mendelian randomization estimate (quantity 3) to estimate the annual total healthcare costs and QALYs saved by reducing incidence of prostate cancer with the intervention.

Because each quantity had associated uncertainty, we repeated the hypothetical intervention 10,000 times, with each quantity taken from a normal distribution with a mean of the effect estimate, and a standard deviation of the standard error of the effect estimate. We present the median of the resulting estimates as the point estimate for the effect of the intervention, with the 2.5 and 97.5 percentiles as the 95% confidence interval. This approach (implicitly) accounts for survival up to a median of seven years of follow up.

We estimated the cost-effectiveness of SGLT2 inhibition for population-wide prophylaxis of incident prostate cancer. Using estimates from our conventional multivariable and Mendelian Randomization estimates of the impact of prostate cancer on healthcare costs and quality-adjusted life years, we compared the impact of SGLT2 inhibition on prostate cancer risk assuming that the cost of doing so was equivalent to the cost 300mg of daily canagliflozin.

We then compared the cost and effect of so doing to a cost-effectiveness threshold of £20,000 using a net monetary benefit calculation. This threshold is routinely used by the National Institute of Health and Care Excellence (NICE) to appraise the value of new medical technologies, including pharmacotherapeutic agents. Under this approach, interventions are cost-effective when their incremental benefit (measured as the difference between intervention benefits and comparator benefits) scaled by the cost-effectiveness threshold exceed incremental costs (measured as the difference between intervention costs and comparator costs). We measure benefits in the present context using quality-adjusted life years, and compared population-wide SGLT2 inhibition to a “do nothing” comparator, which reflects the absence of any accepted pharmacotherapeutic intervention for reducing population-wide prostate cancer risk.

## 3 Data

### 3.1 Study population

UK Biobank is a population-based health research resource consisting of over 500,000 people, who were recruited between the years 2006 and 2010 from 22 centres across the UK (6). We used medical data from hospital episode statistics (HES) linked to all participants up to 31^st^ March 2015, and primary care (general practice) data linked to approximately 31% of UK Biobank participants registered with GP surgeries using EMIS Health (EMIS Web) and TPP (SystmOne) software systems, up to 31^st^ March 2017. The study design, participants and quality control methods have been described in detail previously (7–9). UK Biobank received ethics approval from the Research Ethics Committee (REC reference for UK Biobank is 11/NW/0382). Further information on genotyping is available in (10).

We restricted the main analysis to unrelated individuals of white British ancestry (to avoid confounding by population stratification) living in England or Wales at recruitment. In our sample, 38% of participants had primary care data.

### 3.2 Identification of cancers and creation of polygenic risk scores

We used UK Biobank as the source of our outcome data, the creation of which is described below. To avoid biases from sample overlap (55), we searched for genome-wide association studies (GWASs) on any type of cancer not using data from UK Biobank cohort. We searched the MR Base (mrbase.org) database of GWASs and the Elizabeth Blackwell Institute GWAS catalogue to identify cancers with at least clumped 2 single nucleotide polymorphisms (SNPs) associated with the cancer.

We also restricted our analysis to cancers with specific international classification of disease (ICD) codes at one decimal place, for example, multiple myeloma is coded as “230.0” in ICD-9, and “C90.0” in ICD-10. Where we found multiple GWAS for the same cancer, we preferentially used the GWAS with the most participants.

For each cancer, we clumped the genome-wide significant SNPs at an R^2^ threshold of 0.001 within a 10,000 kilobase window. We searched for proxies for all SNPs not in UK Biobank using the European subsample of 1,000 genomes as a reference panel (with a lower R^2^ limit of 0.6) (11). We used all included SNPs to construct PRS for each cancer, calculated as the weighted sum of the SNP effect alleles for all SNPs associated with each cancer, with each SNP weighted by the regression coefficient from the corresponding GWAS.

For some cancers, we found multiple GWASs which had subgroups of cancer as the outcome. In these cases, we created PRSs for the main cancer (e.g. breast cancer) as well as the available subgroups (e.g. ER+ and ER-breast cancers). There was insufficient detail in the ICD codes to determine the subgroup of cancer for all participants in UK Biobank, and as consequence the exposure used in the main analyses and reported below was for the general (e.g. breast cancer) exposure rather than its subtypes. In Supplementary Table S3 we also report Mendelian Randomization analysis using the available cancer subgroups.

Following this process, the following cancers were available to be analysed: bladder, breast, colorectal, lung, multiple myeloma, ovarian, prostate and thyroid. Box 1 contains details for all PRS we generated for these cancers, including the source GWAS, the number of SNPs in the PRS, and the R^2^ value of the PRS and the cancer from logistic regression with no covariates. Supplementary Table 1 contains details for all SNPs used these PRSs. We created PRS for 8 cancers that had suitable GWAS, with an additional 13 PRS for subgroups of these cancers (2 for breast cancer, 2 for lung cancer, 9 for ovarian cancer), see Box 1.

#### Box 1

Creation of cancer polygenic risk scores

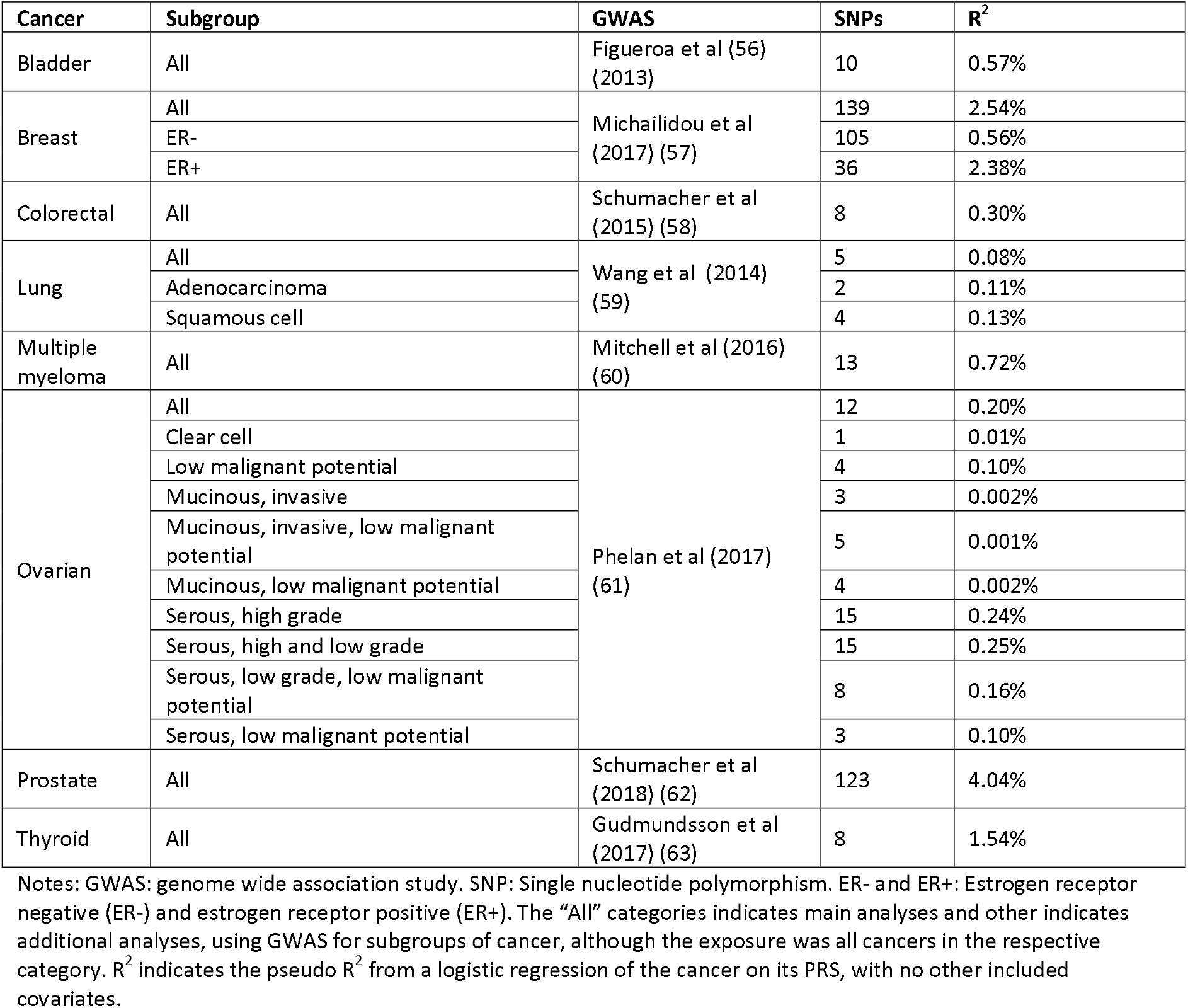

We used age, sex and UK Biobank recruitment centre reported at the UK Biobank baseline assessment as covariables, as well as 40 genetic principal components derived by UK Biobank to control for population stratification (35).

### 3.3 Estimation of quality-adjusted life years

We created health-related quality of life for all participants in UK Biobank from recruitment until March 2017. The details of how these data were constructed are given in Harrison et al (64). In brief, we used the data on quality of life in Sullivan et al (65) for 240 health conditions to create person-specific quality of life scores for every individual for all participants per day in UK Biobank from recruitment to 31 March 2017 or death, whichever came first. We multiplied the results for QALYs by 100 to give the percentage of a QALY changed by having versus not having the cancer. Note that the QALY estimates represent the change in QALYs over an average year of follow-up; that is, they are not cumulative over time.

These health conditions were recorded in primary care records, hospital records (hospital episode statistics) or self-reported in the baseline UK Biobank recruitment questionnaire. These data were averaged over years of follow-up to create quality-adjusted life years (QALYs) for each individual. The full list of health conditions used to create these QALYs is reported in Supplementary Table 2, alongside the ICD-9, ICD-10, Read version 2 and 3 codes (for primary care) associated with each condition.

### 3.4 Estimation of healthcare costs

We have previously reported our approach to creating healthcare cost for the UK Biobank cohort (22, 66). Briefly, we created healthcare costs from records of procedure and diagnosis codes recorded in Hospital Episode Statistics linked to the UK Biobank cohort. This encompassed all inpatient care (for which patients occupy a bed but do not necessarily stay overnight) for publicly funded care in NHS hospitals from recruitment into UK Biobank to 31 March 2015. We used these data to calculate inpatient hospital costs per-patient per year of follow-up. Primary care costs from recruitment into UK Biobank until 31 March 2017 were estimated from the costs of primary care consultations and from the cost of prescribed drugs (as of November 2019). Further details on the construction of primary care costs are given in Harrison et al (64).

We combined annual primary and secondary care healthcare costs to create a total annual healthcare cost figure per person for each year of follow up. This comprised costs related to inpatient hospital care episodes, primary care appointments and primary care drug prescriptions. Primary care data were available for 38% of the sample. We used multiple imputation by chained equations to impute primary care healthcare costs and other outcomes where data were missing. We created 100 imputed datasets (16) and analysed these datasets using Rubin’s rules. Other elements of care, such as emergency care and private healthcare undertaken in private facilities, were not available as outcomes for this cohort. Including these costs would likely increase the overall size of our total cost estimate per person but is very unlikely to alter the magnitude of the associations we estimate in conventional or Mendelian Randomization models.

The price for 300mg of canagliflozin, used as an exemplar SGLT2 inhibitor to model the cost-effectiveness of offering a population wide intervention to reduce prostate cancer risk, was taken from the 2019 NHS drug tariff price (67). The monthly price for 30 canagliflozin tablets of £39.20 was grossed up to give an annual cost figure of £470.

All costs used in our analysis were expressed in 2019 prices. Cost data not expressed in 2019 prices were inflated to this price level using the NHS cost inflation index (68).

### 3.5 Data and code availability

The empirical dataset will be archived with UK Biobank and made available to individuals who obtain the necessary permissions from the study’s data access committees. The code used to clean and analyse the data is available here: https://github.com/sean-harrison-bristol/

## 4 Results

### 4.1 Introduction

We analysed data on 310,913 unrelated individuals, of whom 166,981 (53.7%) were female. Mean age at recruitment was 56.9 years (standard deviation: 8 years), and participants were followed up for a mean of 8.1 years (standard deviation 0.8 years). Median annual total healthcare costs per person were £601 (interquartile range: £212 to £1,217). The most prevalent cancer we examined in UK Biobank was breast cancer (6.6% of women), followed by prostate cancer (4.3% of men), colorectal cancer (1.4%), bladder cancer (0.7%), lung cancer (0.7%), ovarian cancer (0.7% of women), multiple myeloma (0.2%), and thyroid cancer (0.2%).

### 4.2 Results of main analysis

Estimates from conventional multivariable analysis indicated that cancer diagnoses reduced annual QALYs per person and increased annual healthcare costs per person (Table 2). The quantitative impacts were material for all cancers; the largest absolute impacts for QALYs arose in relation to lung cancer and multiple myeloma for healthcare costs.

**Table 1.**
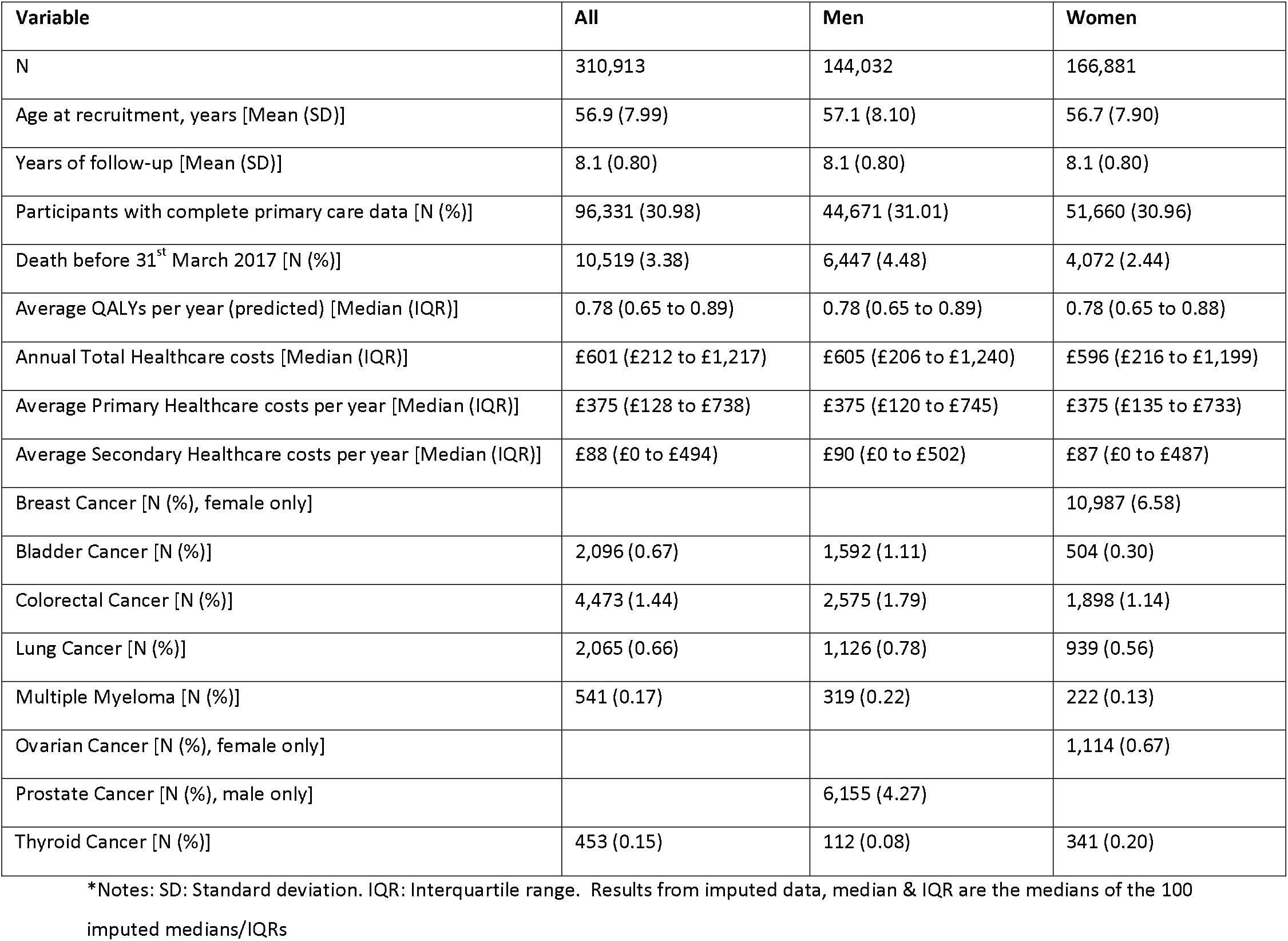
Summary of analysis sample

| Variable | All | Men | Women |
| --- | --- | --- | --- |
| N | 310,913 | 144,032 | 166,881 |
| Age at recruitment, years [Mean (SD)] | 56.9 (7.99) | 57.1 (8.10) | 56.7 (7.90) |
| Years of follow-up [Mean (SD)] | 8.1 (0.80) | 8.1 (0.80) | 8.1 (0.80) |
| Participants with complete primary care data [N (%)] | 96,331 (30.98) | 44,671 (31.01) | 51,660 (30.96) |
| Death before 31<sup>st</sup> March 2017 [N (%)] | 10,519 (3.38) | 6,447 (4.48) | 4,072 (2.44) |
| Average QALYs per year (predicted) [Median (IQR)] | 0.78 (0.65 to 0.89) | 0.78 (0.65 to 0.89) | 0.78 (0.65 to 0.88) |
| Annual Total Healthcare costs [Median (IQR)] | £601 (£212 to £1,217) | £605 (£206 to £1,240) | £596 (£216 to £1,199) |
| Average Primary Healthcare costs per year [Median (IQR)] | £375 (£128 to £738) | £375 (£120 to £745) | £375 (£135 to £733) |
| Average Secondary Healthcare costs per year [Median (IQR)] | £88 (£0 to £494) | £90 (£0 to £502) | £87 (£0 to £487) |
| Breast Cancer [N (%), female only] |  |  | 10,987 (6.58) |
| Bladder Cancer [N (%)] | 2,096 (0.67) | 1,592 (1.11) | 504 (0.30) |
| Colorectal Cancer [N (%)] | 4,473 (1.44) | 2,575 (1.79) | 1,898 (1.14) |
| Lung Cancer [N (%)] | 2,065 (0.66) | 1,126 (0.78) | 939 (0.56) |
| Multiple Myeloma [N (%)] | 541 (0.17) | 319 (0.22) | 222 (0.13) |
| Ovarian Cancer [N (%), female only] |  |  | 1,114 (0.67) |
| Prostate Cancer [N (%), male only] |  | 6,155 (4.27) |  |
| Thyroid Cancer [N (%)] | 453 (0.15) | 112 (0.08) | 341 (0.20) |

\*Notes: SD: Standard deviation. IQR: Interquartile range. Results from imputed data, median & IQR are the medians of the 100 imputed medians/IQRs

**Table 2:** Results from the multivariable adjusted analyses

| Exposure | Outcome | Beta (95% CI) |
| --- | --- | --- |
| Bladder Cancer | QALYs per year | -12.42% (-13.37% to -11.47%) |
|  | Total healthcare costs per year | £1,346 (£1,282 to £1,410) |
| Breast Cancer | QALYs per year | -4.73% (-5.14% to -4.31%) |
|  | Total healthcare costs per year | £704 (£676 to £732) |
| Colorectal Cancer | QALYs per year | -11.20% (-11.87% to -10.53%) |
|  | Total healthcare costs per year | £1,402 (£1,358 to £1,446) |
| Lung Cancer | QALYs per year | -25.89% (-26.87% to -24.91%) |
|  | Total healthcare costs per year | £1,564 (£1,499 to £1,629) |
| Multiple Myeloma | QALYs per year | -18.06% (-19.90% to -16.22%) |
|  | Total healthcare costs per year | £3,488 (£3,363 to £3,612) |
| Ovarian Cancer | QALYs per year | -11.04% (-12.28% to -9.79%) |
|  | Total healthcare costs per year | £1,431 (£1,347 to £1,516) |
| Prostate Cancer | QALYs per year | -4.89% (-5.45% to -4.32%) |
|  | Total healthcare costs per year | £552 (£512 to £592) |
| Thyroid Cancer | QALYs per year | -5.20% (-7.19% to -3.22%) |
|  | Total healthcare costs per year | £686 (£549 to £822) |

Notes: CI: confidence interval; QALY: quality-adjusted life year.

Table 3 presents the Mendelian Randomization estimates for the same cancer exposures and outcomes. The estimates were relatively precise for breast cancer, showing a clear healthcare cost and reduction in QALYs (£798 per year, 95% CI: £549 to £1,048 and -5.51% QALYs per year, 95% CI: -9.32% to -1.70%), and for prostate cancer, which had much lower healthcare costs and change in QALYs (£134 per year, 95% CI: -£217 to £485 per year, and -2.68% QALYS per year, 95% CI: -7.48% to 2.12%).

**Table 3:** Results from the Mendelian randomization analyses

| Exposure | Outcome | Beta (95% CI) |
| --- | --- | --- |
| Bladder Cancer | QALYs per year | 1.83% (-42.23% to 45.88%) |
|  | Total healthcare costs per year | £445 (£-2,569 to £3,459) |
| Breast Cancer | QALYs per year | -5.51% (-9.32% to -1.70%) |
|  | Total healthcare costs per year | £798 (£549 to £1,048) |
| Colorectal Cancer | QALYs per year | -21.38% (-51.70% to 8.94%) |
|  | Total healthcare costs per year | £1,917 (£-181 to £4,016) |
| Lung Cancer | QALYs per year | -49.39% (-163.32% to 64.55%) |
|  | Total healthcare costs per year | £2,732 (£-5,045 to £10,508) |
| Multiple Myeloma | QALYs per year | -80.90% (-220.04% to 58.25%) |
|  | Total healthcare costs per year | £10,720 (£1,308 to £20,133) |
| Ovarian Cancer | QALYs per year | -100.16% (-213.00% to 12.68%) |
|  | Total healthcare costs per year | £4,779 (£-2,289 to £11,847) |
| Prostate Cancer | QALYs per year | -2.68% (-7.48% to 2.12%) |
|  | Total healthcare costs per year | £134 (£-217 to £485) |
| Thyroid Cancer | QALYs per year | -94.41% (-207.80% to 18.98%) |
|  | Total healthcare costs per year | £12,071 (£4,332 to £19,810) |

Notes: CI: confidence interval; QALY: quality-adjusted life year.

Estimates were imprecise for all other cancers. The Mendelian randomization estimates are necessarily less precise than the corresponding conventional estimates, since their precision is a function of the amount of variance in each cancer exposure explained by the respective polygenic risk score.

F statistics were lowest for lung (F-statistic = 21) and ovarian (F-statistic = 25) cancers, suggesting possible weak instrument bias, but very high for prostate (F-statistic = 1,727) and breast (F-statistic = 1,824) cancers, see Supplementary table S4. This suggests little evidence for weak instrument bias for those exposures for which the most precise estimates were obtained in the main analysis.

The multivariable adjusted analyses were generally consistent with the Mendelian randomization analyses, with relatively little evidence from endogeneity tests for differences (except for thyroid cancer), see Supplementary Table S6. Large cost impacts were apparent for multiple myeloma and thyroid cancer which differed drastically from the multivariable adjusted estimates for these cancers. Although no evidence of horizontal pleiotropy was observed for either exposure (see Supplementary Table S3), it is possible that genetic variants for both cancers indicate such large cost impacts because they may be associated with a heritable factor that is upstream of each cancer exposure that itself increases costs.

### 4.3 Mendelian Randomization sensitivity analysis

We found little evidence of pleiotropy in the Mendelian Randomization estimates for breast and prostate cancer, suggesting little evidence that the exclusion restriction was violated. The p-values for SNP heterogeneity were between 0.59 and 1.00 for Cochran’s Q statistic, and estimates from IVW, Egger, penalized weighted median, and weighted mode Mendelian Randomization estimates were all broadly similar, see Supplementary Table S3. There was little evidence of pleiotropy for the other exposures, although the number of SNPS available for analysis for these other cancers was smaller than for prostate and breast cancer.

We examined results stratified by sex (Supplementary Table S4). However, as the most precise estimates were for breast and prostate cancer (each affecting predominantly one sex), there was little evidence of differences between sexes for any other cancer. We also examined results split by both age and diagnosis time (pre- or post-baseline) see Supplementary Table S4. For breast cancer, the effect of breast cancer on QALYs per year was higher but more imprecise for both cancers diagnosed pre-baseline and post-baseline, whereas total healthcare costs per year were highest for cancers diagnosed post-baseline (Mendelian randomization estimate: £2,300 per year, 95% CI: £1,575 to £3,026).

For prostate cancer, there was little difference in the effects on healthcare costs and QALYs per year when split by either diagnosis time or age. There was little difference in the effects on healthcare costs and QALYs per year when split by age.

### 4.4 Evaluating the impact of SGLT2 inhibition on prostate cancer risk

We used the Mendelian Randomization estimates of the effect of genetic liability to prostate cancer on healthcare costs and QALYs to model the impact of SGLT2 inhibition. Our hypothetical intervention assumed that every man in the analysis sample was offered daily SGLT2 inhibitors from the start of follow-up in UK Biobank. Since there are no population-wide prophylactic pharmacotherapeutic agents which are known to reduce prostate cancer incidence, we assessed these effects against a do nothing” comparator. Given odds ratios for incidence of prostate cancer of 0.29 (95% CI=0.13 to 0.65) and 0.51 (95% CI: 0.33 to 0.79) drawn from Zheng et al (52), we then applied our new conventional multivariable and Mendelian Randomization estimates, accounting for associated uncertainty, of the effect of genetic liability to prostate cancer on healthcare costs and QALYs to model cost-effectiveness of population wide SGLT2 inhibition in this cohort of men against a “do-nothing” comparator.

Results for the analyses using an OR of 0.29 (95% CI=0.13 to 0.65) for the intervention for total healthcare costs and QALYs for different age groups are shown in Table 4. As a sensitivity analysis, we also repeated this analysis assuming the more conservative OR (0.51, 95% CI: 0.33 to 0.79 .of the replication analysis in Zheng et al (52) and report our findings in Supplementary Table S5.

**Table 4:** Results from the SGLT2 intervention analysis compared to a “do nothing” comparator

| Age group | Outcome | Analysis Method | Effect estimate (95% CI) |
| --- | --- | --- | --- |
| All | Annual total healthcare costs | Mendelian randomisation | £463.45 (£444.11 to £481.33) |
|  |  | Multivariable adjusted | £451.47 (£446.71 to £460.80) |
|  | QALYs per year | Mendelian randomisation | 0.13% (-0.1% to 0.4%) |
|  |  | Multivariable adjusted | 0.09% (0.04% to 0.13%) |
|  | Net monetary benefit at cost-effectiveness threshold of £20,000 | Mendelian randomisation | -£438 (-£494 to -£372) |
|  |  | Multivariable adjusted | -£433 (-£452 to -£422) |
| <50 years | Annual total healthcare costs | Mendelian randomisation | £477.28 (£459.55 to £497.65) |
|  |  | Multivariable adjusted | £468.75 (£467.81 to £469.55) |
|  | QALYs per year | Mendelian randomisation | -0.06% (-0.28% to 0.15%) |
|  |  | Multivariable adjusted | 0% (-0.01% to 0.02%) |
|  | Net monetary benefit at cost-effectiveness threshold of £20,000 | Mendelian randomisation | -£488 (-£545 to -£438) |
|  |  | Multivariable adjusted | -£468 (-£470 to -£465) |
| 50 to 59 years | Annual total healthcare costs | Mendelian randomisation | £468.29 (£446.21 to £490.54) |
|  |  | Multivariable adjusted | £459.01 (£455.51 to £464.69) |
|  | QALYs per year | Mendelian randomisation | 0.02% (-0.3% to 0.37%) |
|  |  | Multivariable adjusted | 0.05% (0.02% to 0.09%) |
|  | Net monetary benefit at cost-effectiveness threshold of £20,000 | Mendelian randomisation | -£462 (-£539 to -£385) |
|  |  | Multivariable adjusted | -£448 (-£460 to -£439) |
| 60+ years | Annual total healthcare costs | Mendelian randomisation | £448.79 (£414.55-£478.39) |
|  |  | Multivariable adjusted | £439.30 (£430.51 to £455.37) |
|  | QALYs per year | Mendelian randomisation | 0.33% (-0.07% to 0.81%) |
|  |  | Multivariable adjusted | 0.15% (0.07% to 0.22%) |
|  | Net monetary benefit at cost-effectiveness threshold of £20,000 | Mendelian randomisation | -£384 (-£477 to -£268) |
|  |  | Multivariable adjusted | -£409 (-£441 to -£388) |

Notes: CI: confidence interval; QALY: quality-adjusted life year.

Under the assumptions of this analysis, the provision of SGLT2 inhibition as a prophylactic population wide intervention dominates the “do nothing” status quo for men at risk of prostate cancer in the sense that it lowers healthcare costs and increases quality adjusted life years, but only if offered at zero cost (see Supplementary Table S6). This does not hold once the costs of the drugs themselves are accounted for (Table 4). Net monetary benefit becomes positive only if the SGLT2 inhibition drugs are offered at very modest prices; for example, under the “All” scenario above, the drug becomes just cost-effective (in the sense of resulting in a positive net monetary benefit when accounting for other costs and the associated quality of life impacts) at an annual cost of approximately £32.

## 5 Discussion

We developed the first Mendelian Randomization estimates of the impact of cancer status on healthcare cost and on quality of life, the two outcomes necessary to undertake a comprehensive economic evaluation of any intervention. We studied the impacts of bladder, breast, colorectal, lung, multiple myeloma, ovarian, prostate and thyroid cancers on these outcomes.

The Mendelian Randomization estimates are less likely to be biased by confounding from omitted variables and reverse causation than conventional analytic study designs such as those based on cohort studies or decision-analytic simulation models using observational effect estimates (4). We used the largest available GWASs to identify cancer exposures without restriction on site. Our outcome data were drawn from the large UK Biobank study which has comprehensive linked data which permitted the calculation of patient-level healthcare costs, quality of life and disease status over many years.

The estimates will be particularly relevant to cost-effectiveness models evaluating the impact of cancer on healthcare costs and on quality of life. The policy evaluation element of our paper provides an example of how these types of estimates may be used to simulate the long-term impacts of anti-cancer intervention in the absence of, or in advance of, a randomized controlled trial. These methods could also be used to prioritize future randomized controlled trials by identifying therapies most likely to be cost-effective.

Adjusted multivariable estimates indicated material impacts of cancer status on healthcare costs and quality of life for all 8 included cancers. Estimates from the Mendelian Randomization models were much more imprecise, although genetic liability to breast and prostate cancers was associated with reductions in QALYs and increases in healthcare cost. There was little evidence for heterogeneity and pleiotropy in violation of the exclusion restriction between SNPs for these cancers. Summary Mendelian randomization sensitivity estimates were similar to each other and broadly consistent with the main PRS estimates, indicating the same causal effect was likely being identified in each model without gross bias from violations of the exclusion restriction.

The results from the Mendelian Randomization analyses on cost and QALY outcomes are local average treatment effect estimates that represent the effect of lifelong exposure to higher germline genetic liability to each respective cancer. The results of the analysis cannot therefore be interpreted as, for example, the impact on costs of reducing cancer risk at any particular timepoint in the life course. Instead, they represent the impact on cost and QALY outcomes from changing long-term germline risk in this population for each specific cancer. They may not correspond to the magnitude of changes in these outcomes that would be observed through some other modification of cancer risk through, for example, manipulation of environmental influences on cancer risk (69).

The cancer risks we modelled reflect the risks of incident disease. Our analysis includes SNPs that may influence both incidence and progression. For some cancers, especially prostate cancer, identifying aggressive cancer is arguably more important than understanding incidence since most tumours in (for example) the prostate do not influence overall survival (70). However, tumours that do not affect survival may still influence cost and quality of life and their influence will be reflected in our estimates. Nevertheless, there would be value in updating this analysis when large GWASs with an explicit focus on cancer progression and aggression are made available.

Healthcare costs were estimated from inpatient care episodes, primary care consultations and primary care drug prescriptions obtained or imputed from electronic primary care records. Follow-up was two years shorter for secondary care costs than primary care costs but was averaged over years of follow-up and should not have affected results. We did not capture all costs, including private care delivered in private healthcare facilities or outpatient care, neither of which were available for this cohort. This suggests our results may underestimate the costs associated with cancer.

Despite its size, UK Biobank is not representative of the UK population as self-selected participants tend to be wealthier and healthier compared to wider population from which it was recruited (31). It is possible, although we cannot demonstrate, that our Mendelian Randomization results have underestimated the costs and quality of life impacts of cancer, since wealthier and healthier people may have more capacity and material resources to ameliorate the detrimental effects on healthcare cost and quality of life of increased genetic liability to cancer. The scale of this type of selection and any associated bias is unknown, but may have less significance for our conclusions than biases arising from violations of the instrumental variable assumptions (71, 72), for which we separately report sensitivity analyses and for more biologically distal exposures (73) than we study. We controlled for genetic principal components in our analysis; there is evidence that this may not control for all remaining population or geographic structure (74, 75) (33), both which could give rise to residual biases.

Although Mendelian randomization is likely to be less affected by confounding and reverse causality than conventional multivariable adjusted analyses, our analysis of a sample of unrelated individuals may give rise to further biases from dynastic effects and assortative mating. Dynastic effects refer to the influence of parental genotype on offspring phenotype. Parents (and both their transmitted and untransmitted alleles) may influence the environment in which their children are raised and could confound the SNP-cancer associations we estimate later in the life of these offspring. Given that health costs are measured in middle to early old age, these impacts are probably modest, if present at all. Assortative mating refers to non-random mating; that is, individuals may preferentially mate (even if inadvertently) with others possessing alleles that increases the risk of cancer. This will tend to create clusters of similar alleles in particular environments and may re-introduce environmental confounding otherwise avoided in Mendelian Randomization.

Within-family Mendelian randomization studies can account for some of these biases. However, we did not have the statistical power to conduct these analyses since the binary cancer exposures we studied affected relatively few people in UK Biobank (76). In any event, there are as yet no large cohort of family trios that have reached middle and early old age that would have enabled this analysis for our UK Biobank cohort, and the magnitude of potential biases may from these sources may not be large for the cancer exposures that we study. For more visible traits such as BMI (77, 78) and biologically distal phenotypes such as education (79) there is robust evidence of for family-related effects assortative mating and dynastic effects (80), but it is less clear if these processes have any significance for the cancer exposures studied here.

We measured the quality of life of UK Biobank participants by assigning quality of life decrements associated with 240 specific health conditions. This permitted the calculation and use of QALYs as an outcome. We note that, amongst these conditions, are some (but not all) of the cancers that we also study as exposures (specifically bladder, breast, colon, lung, and prostate – see Supplementary Table S2). This creates a modest degree of circularity, but we don’t consider this as problematic because the cancers we study contribute relatively little variance in QALYs across the full sample given their prevalence, although this variation is still sufficient to identify the causal effects of exposure to these cancer in the Mendelian Randomization analysis. Moreover, these Mendelian Randomization estimates should also in principle account for any other health conditions downstream of the specific cancer exposures we study and which would otherwise be omitted from conventional multivariable analysis.

We used both conventional multivariable and Mendelian Randomization estimates to assess the potential cost-effectiveness of a hypothetical population-wide prophylactic intervention for prostate cancer based on SGLT2 inhibition. Assuming the costs of this intervention would be as for 300mg canagliflozin, this intervention was unlikely to be cost-effective at this price under the assumptions of that analysis which included (but were not limited) to reductions in the odds of prostate cancer based on the findings of Zheng et al (52), and the absence of side effects.

Even given these limitations, we consider that these analyses demonstrate at least two important conclusions. The first is to lend support for the conduct of a randomized controlled trial of SGLT2 inhibitors. The work of Zheng et al (52) indicated that they may have substantial impacts on the odds of prostate cancer, and are the sole pharmacotherapy to demonstrate such an impact as a prophylactic for incident prostate cancer. Although the present work demonstrates that such an intervention is unlikely to be cost-effective, this conclusion depends on the price at which the rights holders of SGLT2 inhibitors would offer the drug for a novel prostate cancer indication.

The second conclusion is that the methods we have developed and applied here for the first time represent a feasible and rapid way of assessing the cost-effectiveness of both novel and established interventions. These methods are an alternative but complementary approach to, for example, Markov cohort simulations and related study designs that seek to model the long-term impact of exposures and interventions on health economic outcomes. We do not consider these methods as an alternative to fully articulated RCTs but they do offer considerable promise as a means of rapidly and efficiently prioritising targets for investigation in subsequent RCTs, provided sufficient genetic evidence for the specific target of interest (see e.g. (81)) exists to enable robust Mendelian Randomization analysis.

## 6 Conclusion

We estimated the causal impact of liability to eight cancers on healthcare costs and on quality-adjusted life years using a Mendelian Randomization study design using data drawn from large genome-wide association studies and from the UK Biobank cohort. We identified impacts from genetic liability to prostate cancer amongst men and breast cancer amongst women on both healthcare costs and quality-adjusted life years. These results were concordant with those obtained from pleiotropy-robust estimators and when stratifying by age and sex. Mendelian Randomization results from less common cancers were imprecise.

We also used both conventional multivariable and Mendelian Randomization results to assess the impact of hypothetical population-wide preventative intervention to reduce the risk of prostate cancer based on the repurposing of SGLT2 inhibitors. Both sets of estimates indicated that this intervention was unlikely to be cost-effective if drugs were priced as for their current anti-diabetes indication.

Robust evidence of the causal impact of cancer exposures on health economic outcomes may be used as inputs into decision analytic models of cancer interventions such as screening programmes or simulations of longer-term outcomes associated with therapies investigated in RCTs with short follow-ups. We also demonstrated a method to rapidly and efficiently estimate the cost-effectiveness of a hypothetical population-scale anti-cancer intervention to inform and complement other means of assessing long-term intervention cost-effectiveness. These estimates and methods will contribute to ongoing debates about managing the economic impacts of cancer in the face of changes in cancer incidence, cancer survivorship, population structure, and ongoing debates about the costs of cancer drug development and the remuneration of anti-cancer therapies.

## Data Availability

The empirical dataset will be archived with UK Biobank and made available to individuals who obtain the necessary permissions from the study's data access committees.

## Acknowledgments

This research was conducted using the UK Biobank Resource under Application Number 29294. We are also very grateful to Jie Zheng for advice on the modelling of SGLT2 inhibition.

